# EHR Foundation Models Improve Robustness in the Presence of Temporal Distribution Shift

**DOI:** 10.1101/2022.04.15.22273900

**Authors:** Lin Lawrence Guo, Ethan Steinberg, Scott Lanyon Fleming, Jose Posada, Joshua Lemmon, Stephen R Pfohl, Nigam Shah, Jason Fries, Lillian Sung

**Author notes:** **ADDRESS FOR CORRESPONDANCE:** Lillian Sung MD, PhD, Division of Haematology/Oncology, The Hospital for Sick Children, 555 University Avenue, Toronto, Ontario, M5G1X8, Canada. co-senior authors (Jason Fries, Lillian Sung).

## Abstract

**Background:** Temporal distribution shift negatively impacts the performance of clinical prediction models over time. Pretraining foundation models using self-supervised learning on electronic health records (EHR) may be effective in acquiring informative global patterns that can improve the robustness of task-specific models.

**Objective:** To evaluate the utility of EHR foundation models in improving the in-distribution (ID) and out-of-distribution (OOD) performance of clinical prediction models.

**Methods:** The cohort consisted of adult inpatients admitted between 2009-2021. Gated recurrent unit (GRU)- and transformer (TRANS)-based foundation models were pretrained on EHR of patients admitted between 2009-2012 and were subsequently used to construct patient representations (CLMBR). These representations were used to learn logistic regression models (CLMBR_GRU_ and CLMBR_TRANS_) to predict hospital mortality, long length of stay, 30-day readmission, and ICU admission. We compared CLMBR_GRU_ and CLMBR_TRANS_ with baseline logistic regression models learned on count-based representations (count-LR) and end-to-end (ETE) GRU and transformer models in ID (2009-2012) and OOD (2013-2021) year groups. Performance was measured using area-under-the-receiver-operating-characteristic curve, area-under-the-precision-recall curve, and absolute calibration error.

**Results:** Models trained on CLMBR generally showed better discrimination relative to count-LR in both ID and OOD year groups. In addition, they often matched or were better than their ETE counterparts. Finally, foundation models’ performance in the self-supervised learning task tracked closely with the ID and OOD performance of the downstream models.

**Conclusions:** These results suggest that pretraining foundation models on electronic health records is a useful approach for developing clinical prediction models that perform well in the presence of temporal distribution shift.

## INTRODUCTION

The large increase in the adoption of electronic health records (EHR) has enabled the use of machine learning to develop highly performant clinical prediction models that have the potential to improve the care of patients[1]. However, the non-stationary healthcare environment can bring about changes in the data distribution between model development and deployment[2], which can degrade the model’s performance over time[3] and consequently its clinical utility[4]. In this study, we explored temporal distribution shift alongside the suitability of *foundation models[5]* – deep neural networks trained on large-scale unlabeled data using self-supervised learning – and whether they can be adapted via transfer learning to improve the robustness of clinical prediction models in the presence of temporal distribution shift.

The cause of temporal distribution shift in clinical medicine is often subtle[6] and the extent of its impact on model performance is heterogeneous across tasks[3, 7-9]. Nonetheless, the consequence of the impact on patient care and physician’s trust can be severe. An example is the widely implemented Epic sepsis model developed on data collected between 2013-2015 that performed below expectation when evaluated at Michigan Medicine on data collected between 2018-2019 and resulted in a large number of spurious alerts[4].

Recent approaches that mitigate the impact of temporal distribution shift on model performance in clinical medicine largely rely on model monitoring and updating policies that do not leverage the entire patient population available[10]. In addition, proactive approaches using domain generalization and adaptation have shown little to no success[3].

To date, few studies have explored learning contextualized patient representations at scale. Findings from domains outside of clinical medicine suggest significant performance[11] and robustness[12, 13] benefits to pretraining foundation models. In this study, we adopt CLMBR,[14] an EHR foundation model pretrained on patient timelines comprised of sequential structured medical codes using autoregressive sequence modeling as the self-supervised learning task. Transfer of the structure learned by CLMBR from the entire patient population to downstream models have demonstrated performance benefits compared to standard baselines including count-based models, especially when the number of patient records was small.

In this study, we evaluated the utility of CLMBR in mitigating the impact of temporal distribution shift on model performance. We hypothesized that the global patterns embedded in CLMBR can be adapted into models that perform better than count-based representations in out-of-distribution (OOD) years. In addition, we characterized the robustness of popular architectures used in clinical settings, namely gated recurrent unit (GRU)[14] and transformers (TRANS)[15, 16]. Further, to understand the contribution of CLMBR, we evaluated both GRU and TRANS end-to-end (ETE) models. Therefore, the objectives were to compare the in-distribution (ID) and OOD performance of CLMBR_GRU_, CLMBR_TRANS_, ETE_GRU_ and ETE_TRANS_ compared to models trained using count-based representations.

## METHODS

### Data Source

We used data from the STAnford medicine Research data Repository (STARR)[17]. Data in STARR are routinely collected in the EHR of Stanford Medical Center, comprised of Stanford Health Care (primarily adult-directed care) and Lucile Packard Children’s Hospital (primarily pediatric-directed care). These data are mapped to the Observational Medical Outcomes Partnership Common Data Model (OMOP-CDM), which facilitates multi-center observational research[18, 19]. The resulting dataset was named STARR-OMOP. This study used de-identified data in which dates were jittered by up to 30 days but were accurate within a patient timeline. Because of de-identification, the requirement for Institutional Review Board approval was waived by Stanford Medical Center.

### Cohort

We included adult patients over the age of 18 with admissions to the inpatient unit between EHR inception (2009) and August 22, 2021. Admissions to the inpatient unit were either direct or transfers from the emergency department. Encounters with clinic visit alone and encounters in which patient death or discharges occurred on the same day of admission were excluded. For patients with multiple admissions, one was randomly selected so that each patient was only represented once in the dataset.

### Outcomes

We defined four clinical outcomes. *Hospital mortality* was defined as a patient death occurring during the hospital stay. Long length of stay (LOS) (*long LOS*) was defined as a hospital admission of seven or more days. Readmission in 30 days (*30-day readmission*) was defined as a readmission to an inpatient unit within 30 days after discharge. Intensive care unit (*ICU)* admission was defined as a patient transfer to the intensive care unit during the hospital admission. Each outcome was considered as a binary classification task where the prediction time (also the index time) was set as 11:59PM on the day of admission for the hospital mortality, long LOS and ICU admission tasks, and 11:59PM on the day of discharge for the 30-day readmission prediction task. For the 30-day readmission task, we removed patients who were re-admitted on the day of discharge, and for the ICU admission task, we removed patients transferred on the day of admission since these events would have occurred before prediction time.

### Patient Representations

EHR data corresponding to a particular patient can be treated as a sequence of days that is ordered by time, *d*_*1*_ *… d*_*N*_, where each day consists of a set of events represented by medical codes such as diagnoses, lab tests, procedures, and medication administrations or prescriptions as examples. In this study, we considered two approaches to construct patient representations over the patient timelines as illustrated in Figure 1: count-based representations and CLMBR.

**Figure 1.**
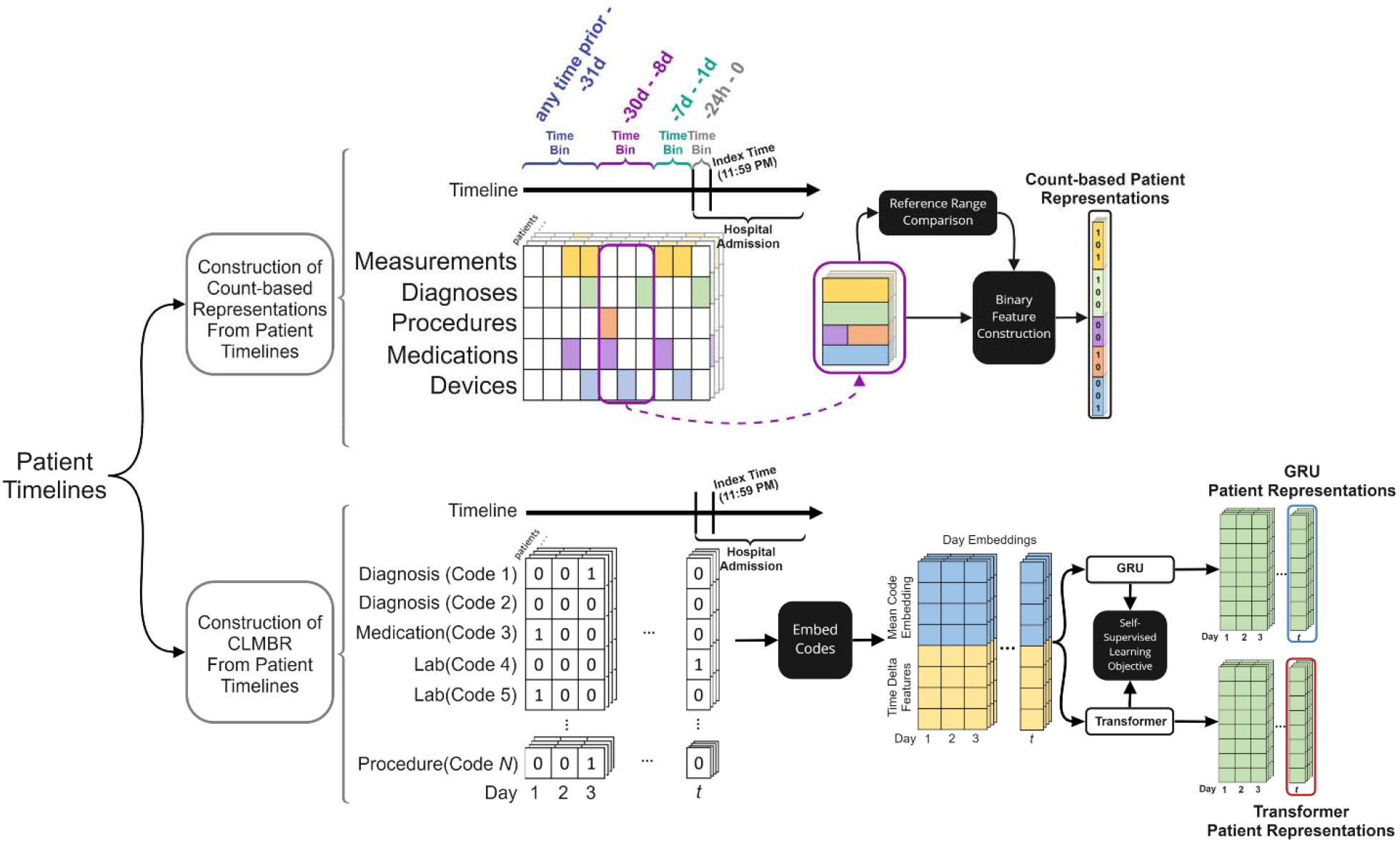
An overview of the two approaches of constructing patient representations used in this study. The purple box in the construction of count-based representations represents the reference range comparison and binary feature construction procedures for a specific time-bin. The construction of CLMBR illustrates the self-supervised pretraining stage, hence the inclusion of the self-supervised learning objective. The construction of CLMBR for the purpose of transfer learning (e.g., for predicting hospital mortality) does not include the self-supervised learning objective. Abbreviations: CLMBR: clinical language model-based representations.

#### Count based representations

The count-based representations were constructed using an open-source count-based featurizer[20] which follows standard practices for patient count-based featurization.[1, 21] This approach constructed patient representations as binary features based on counts of both unique OMOP CDM concepts and derived elements recorded prior to the time of prediction. The feature set consisted of demographic and clinical features.

Demographic features included sex at birth, race, ethnicity, and age at admission discretized into five-year intervals. Clinical features were constructed as the concatenation of the results of a time-dependent extraction procedure applied independently to data elements recorded in time bins defined relative to the time of prediction. The time bins were as follows: 24 hours prior, 1-7 days prior, 8-30 days prior, and 31 days-any time prior. The time-dependent extraction procedure identified all unique OMOP CDM concepts from the following OMOP CDM tables: condition occurrence (diagnosis codes), drug exposure (administration or prescription of medications), procedure occurrence, measurement (includes laboratory tests), device exposure (exposure to implantable objects, medical equipment, supplies, and instruments) and observation (non-standardized tests or clinical observations). Continuous measurement results were represented as binary indicators for abnormal results for each measurement on the basis of whether the result was above or below the reference range.

*Clinical language model-based representations – CLMBR*: The core idea behind CLMBR is that if a sequence model is able to predict sets of medical codes over a patient timeline, then that model may have discovered informative global patterns that can be re-used in various other clinical prediction tasks. Note that the term “language model” in CLMBR merely reflects the similarity in the computations involved between sequence modeling of medical codes and language modeling, and therefore does not indicate natural language processing of any kind.

First, we mapped clinical codes for labs, medications, diagnoses, and procedures to a finite vocabulary of discrete symbols. This vocabulary was then mapped into a clinical ontology to reduce sparsity and used to construct patient sequences for the CLMBR encoder. The medical codes were obtained from the same OMOP CDM tables as used for count-based representations except for the observation table. The Unified Medical Language System (UMLS)[22] was used to extend each medical code to the set of parents in its native ontology when applicable (ICD10 for diagnoses, CPT or MTHH for procedures, and ATC for medications). For instance, the occurrence of the ICD10 code “H61.23” for the diagnosis of impacted cerumen, bilateral, resulted in two additional parent codes, namely “H61.2” (impacted cerumen) and “H61” (other disorders of external ear).

We chose GRU and transformer as the architectures for our sequence models as they have each demonstrated success in the sequence modeling of medical codes in the EHR[14, 18, 19, 23]. To construct patient representations, sets of codes for each day in the patient timeline were first passed through the embedding bag layer of the networks, which computes the mean code embedding for each day. Next, each mean embedding was concatenated with a vector that captured time information including the patient’s age on that day, the time delta from the previous day, whether that day was the first day of the sequence, and the log transform of the age and time delta. Patient representations were then computed by feeding the concatenated vectors into the GRU or transformer (see Supplementary Methods for details on architecture), followed by a linear layer with output size equal to the number of dimensions of the patient representation, which was set to 800 in this study.

To predict the set of codes for a given day, *d*_*i*_, the patient representation from the previous day, *d*_*i-1*_, was used. We formulated the set prediction problem as a series of independent binary classification problems, where the probability of a given code was computed via a sigmoid transformation of the dot product between the code embedding and the patient representation. To deal with the computational complexity of the matrix product induced by the large code space, we used a variant of the hierarchical softmax optimization[24] in which we replaced the softmax transformations with sigmoid transformations. The hierarchical structures of the code space were the same as the ones used for ontology extension (e.g., the hierarchical structure in the ICD10 vocabulary for ICD10 codes). We used the binary cross entropy loss as the loss function during training.

Once the sequence models were trained, we used them to construct representations (the output of the linear layer) for each patient in the cohort to be used by downstream models for clinical prediction tasks (CLMBR_GRU_ and CLMBR_TRANS_). For hospital mortality, long LOS, and ICU admissions, patient representations were obtained up until the day of admission, whereas for 30-day readmission patient representations were obtained up until the day of discharge.

### Experimental Setup

First, we established baseline model performance for each of the four clinical prediction tasks and investigated whether model performance degraded over time as a result of temporal distribution shift. We trained logistic regression models on count-based representations constructed for patients admitted between 2009-2012 (count-LR) and evaluated the models on all years from 2009-2021. The years on which the models were trained (2009-2012) constituted the ID years and the subsequent years (2013-2021) constituted the OOD years for the baseline experiment. We also included oracle models that were trained and evaluated on each of the OOD years for comparison.

Next, we compared ID and OOD performance for four different representation construction and modeling approaches relative to count-LR, namely CLMBR_GRU_, CLMBR_TRANS_, ETE_GRU_, and ETE_TRANS_. For this experiment, ID years were 2009-2012 and the OOD years were 2013-2016 and 2017-2021. For statistical comparisons, we focused on comparing CLMBR_GRU_and CLMBR_TRANS_ vs. count-LR, CLMBR_GRU_ and CLMBR_TRANS_ vs. their respective ETE architecture, and CLMBR_GRU_ vs. CLMBR_TRANS_. To gain insight into relative performance, we subtracted the model’s OOD performance in 2017-2021 by its ID performance in 2009-2012 for the same five representation construction and modeling approaches. To limit multiple testing, describing relative OOD performance and OOD statistical comparisons only focused on 2017-2021 and not 2013-2016.

We also examined the contribution of CLMBR to downstream model performance by examining the Pearson correlation between each sequence model’s pretraining performance and the downstream logistic regression performance in each clinical prediction task. Performance was measured using binary cross-entropy loss.

As a sensitivity analysis, we trained and compared task-specific models on count-based representations and CLMBR using light gradient-boosted machines (LightGBM) instead of logistic regression. In addition, to aid clinical interpretation of the changes in performance between count-LR and CLMBR-based models, we quantified the numbers of decisions that would have been affected if the CLMBR-based models were used instead of count-LR for tasks in which performance degraded over time. Specifically, we selected the better performing CLMBR model (CLMBR_GRU_ vs CLMBR_TRANS_) and calculated the proportion of patients that would have been classified correctly and incorrectly with CLMBR-based models instead of count-LR across various risk thresholds.

#### Model development

The cohort was divided into training (70%), validation (15%), and test (15%) sets with random sampling stratified by the year in which the admission occurred. We extracted count-based representations and CLMBR for each patient admission. For count-based representations, we additionally pruned features with less than 25 observations in the training set. We then pruned the same features from the validation and test sets. To tune GRU and transformer for CLMBR, we performed grid search over the hyperparameter settings for each architecture separately. For GRU, the hyperparameters consisted of learning rate, L2 regularization strength and dropout rate. For transformer, we additionally tuned the number of transformer layers and the rate for code dropout. Supplementary Methods detail the hyperparameter grid and the selected hyperparameter settings. We trained GRUs and transformers on the timelines of 80% of the patients in the training set of 2009-2012 (29026 patients, ∼1.1 million patient days, and ∼41.5 million medical codes), and selected hyperparameter settings based on model performance in the left out 20% of the training set. Then, using the selected GRU and transformer, we constructed CLMBR_GRU_ and CLMBR_TRANS_ for each patient in the cohort.

After computing the representations, we trained logistic regression with L2-regularization and LightGBM models on count-based representations and CLMBR for each clinical outcome in the training set of 2009-2012. Hyperparameter tuning was done on L2 strength, which ranged from 10^−6^ to 10^2^ in increments of powers of 10. We selected hyperparameter values based on the model’s binary cross entropy loss in the validation set of 2009-2012. The ETE models were trained for each clinical outcome separately on the training set of 2009-2012, and hyperparameter tuning was conducted using the same grid as CLMBR_GRU_ and CLMBR_TRANS_ for fair comparisons. We selected hyperparameter settings for ETE models based on model performance in the validation set of 2009-2012. Oracle models for each OOD year (2013-2021) as comparisons for count-LR were trained on count-based representations in the training set of the OOD year, and hyperparameters were selected based on performance in the validation set of the OOD year. Supplementary Methods provide details on the selected hyperparameter setting for logistic regression and ETE models for each clinical prediction task.

CLMBR and ETE models were implemented using Pytorch[25] and were trained on two Nvidia V100 GPUs. We used the Sci-kit Learn’s[26] implementation of logistic regression. Analyses were implemented in Python 3.8[27].

#### Model evaluation

We evaluated each model’s discrimination performance in the test sets using the area-under-the-receiver-operating-characteristic curve (AUROC) and the calibrated area-under-the-precision-recall curve (AUPRC)[28]. The calibrated AUPRC computes the precision using a reference outcome prevalence, here set as the prevalence in the ID year group 2009-2012. Thus, the calibrated AUPRC is invariant to change in outcome prevalence in OOD years and allows us to better interpret its variation over time. We used the absolute calibration error (ACE)[20] as a measure of calibration. ACE is similar to the integrated calibration index[29] but applies a logistic regression estimator to the logit of the predicted probability outputs rather than locally weighted least squares and is thus more computationally efficient.

#### Statistical Analysis

For each metric, we computed the median and 95% confidence interval (CI) of the distribution over performance in the test set obtained from 1000 bootstrap samples. To compare models, we computed the 95% CI of the differences between a pair of models over 1000 bootstrap samples. Statistical significance was defined as comparisons where the 95% CI did not cross 0.

## RESULTS

Supplementary Table 1 presents cohort characteristics for each year and outcome prevalence. Figure 2 shows the impact of temporal distribution shift on performance (AUROC, AUPRC, and ACE) of count-LR trained on admissions from 2009-2012. Model degradation occurred in the OOD years (2013-2021) for long LOS and ICU admission prediction tasks, with larger degradations observed in 2017-2021.

**Figure 2.**
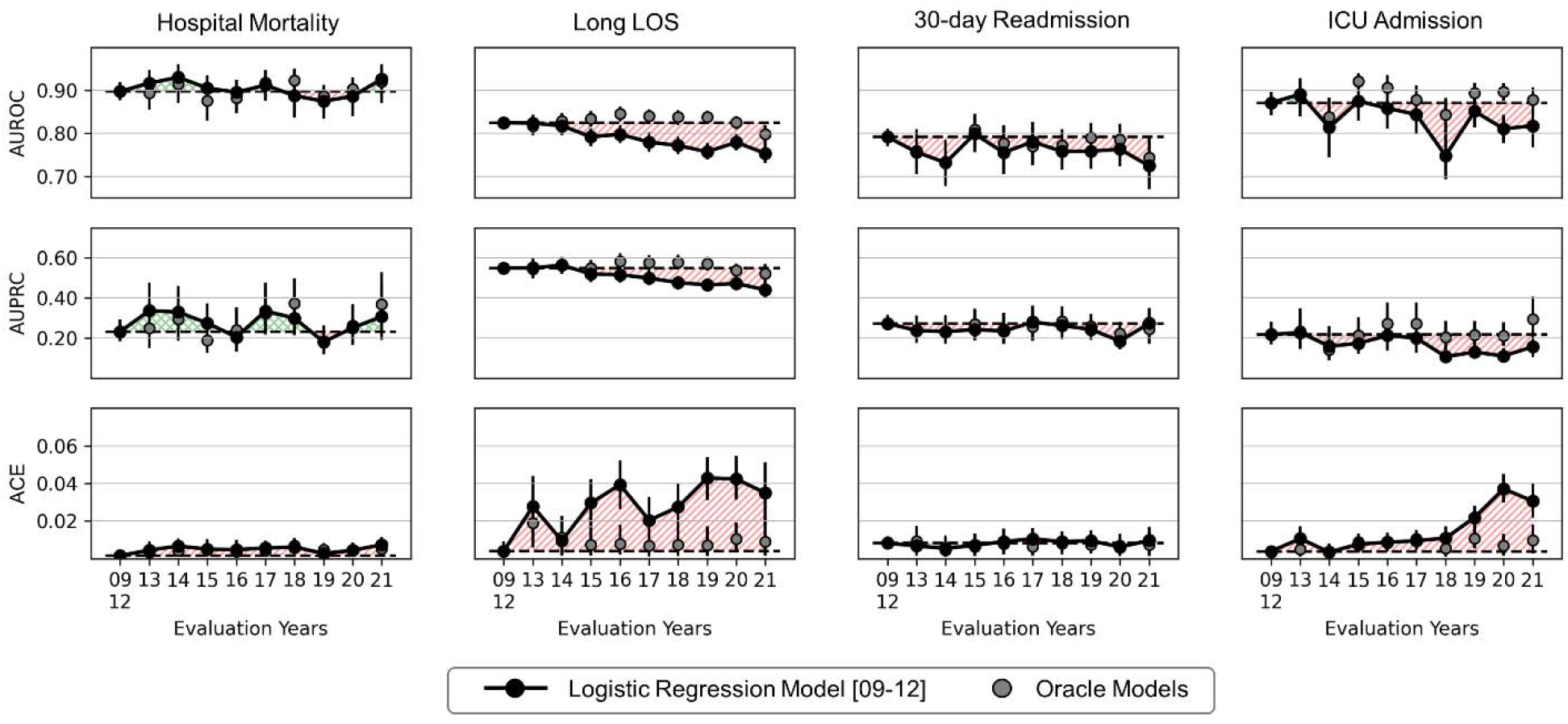
The impact of temporal distribution shift on the performance (AUROC, AUPRC, and ACE) of logistic regression models trained on count-based representations. Shaded regions indicate time windows in which performance in out-of-distribution years (2013-2021) is worse (red) or better (green) than performance in the in-distribution year group (2009-2012). Oracle models were trained and evaluated on each of the out-of-distribution years. Error bars indicate 95% confidence interval obtained from 1000 bootstrap iterations. Abbreviations: AUROC: area under the receiver operating characteristics curve; AUPRC: area under the precision recall curve; ACE: absolute calibration error; LOS: length of stay; ICU: intensive care unit.

Figure 3 shows the relative performance of CLMBR-based and ETE models compared to count-LR in ID (2009-2012) and OOD (2013-2016 and 2017-2021) year groups (see Supplementary Tables 2 and 3 for statistical results of pre-specified comparisons). First, CLMBR_GRU_ and CLMBR_TRANS_ outperformed count-LR in discrimination performance in both ID and OOD year groups across all tasks except for 30-day readmission. In terms of calibration, ID improvement was observed for 30-day readmission for CLMBR_TRANS_ vs. count-LR but other comparisons between CLMBR_GRU_ or CLMBR_TRANS_ vs. count-LR did not show significant differences. In contrast, OOD (2017-2021) calibration results showed deterioration in hospital mortality and 30 day readmission tasks. Second, in general, the ID performance of CLMBR_GRU_ and CLMBR_TRANS_ were similar to or better than ETE_GRU_ and ETE_TRANS_ respectively. However, differences in OOD 2017-2021 performance varied across tasks: CLMBR models generally performed better in predicting ICU admission, and the ETE models generally performed better in predicting long LOS. Third, CLMBR_GRU_ outperformed CLMBR_TRANS_ in discrimination performance in tasks other than 30-day readmission prediction for ID and for OOD hospital mortality and long LOS tasks.

**Figure 3.**
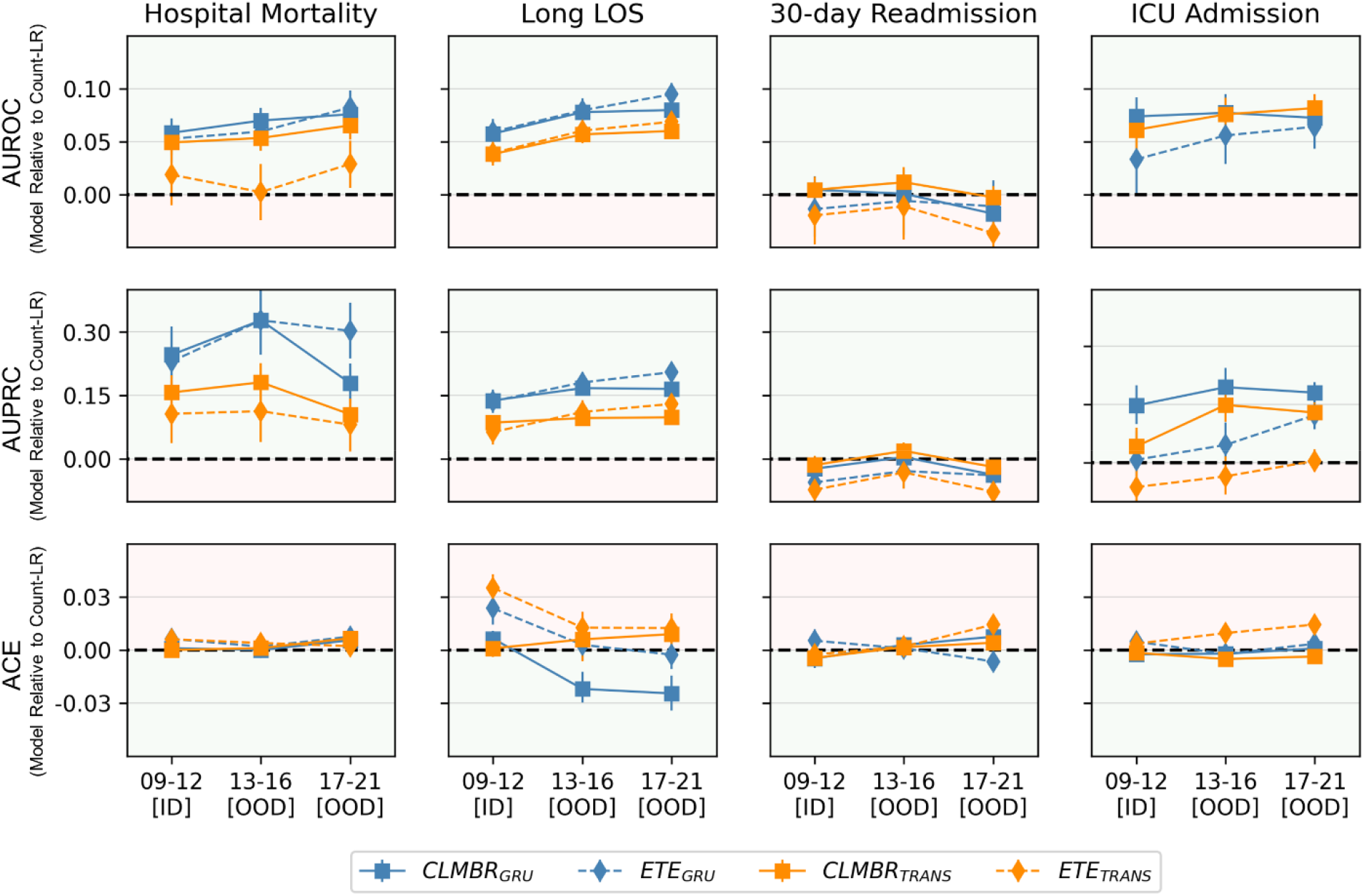
Change in performance (AUROC, AUPRC, and ACE) across representation construction and modeling approaches relative to count-based logistic regression models (Count-LR) in 2009-2012 (09-12), 2013-2016 (13-16), and 2017-2021 (17-21). Performance of count-LR was subtracted to obtain relative performance. Green regions indicate the range of values that is better with respect to count-LR, and red regions indicate the range of values that is worse. Error bars indicate 95% confidence interval obtained from 1000 bootstrap iterations. Supplementary Tables 2 and 3 detail the statistical results. Abbreviations: AUROC: area under the receiver operating characteristics curve; AUPRC: area under the precision recall curve; ACE: absolute calibration error; LOS: length of stay; ICU: intensive care unit; CLMBR: clinical language model-based representation; GRU: gated recurrent unit; Trans: transformer; LR: logistic regression; ETE: end-to-end.

Figure 4 shows relative OOD performance compared to ID performance across representation construction and modeling approaches (see Supplementary Table 4 for statistical results). Comparison of CLMBR_GRU_ or CLMBR_TRANS_ vs. count-LR showed heterogeneous results. In terms of relative discrimination, both CLMBR_GRU_ and CLMBR_TRANS_ had significantly better relative AUROC vs. count-LR for long LOS but were similar for other tasks. In terms of relative calibration, CLMBR_GRU_ and CLMBR_TRANS_ were significantly worse than count-LR for hospital mortality and 30-day readmission tasks. Other comparisons of relative performance were heterogeneous.

**Figure 4.**
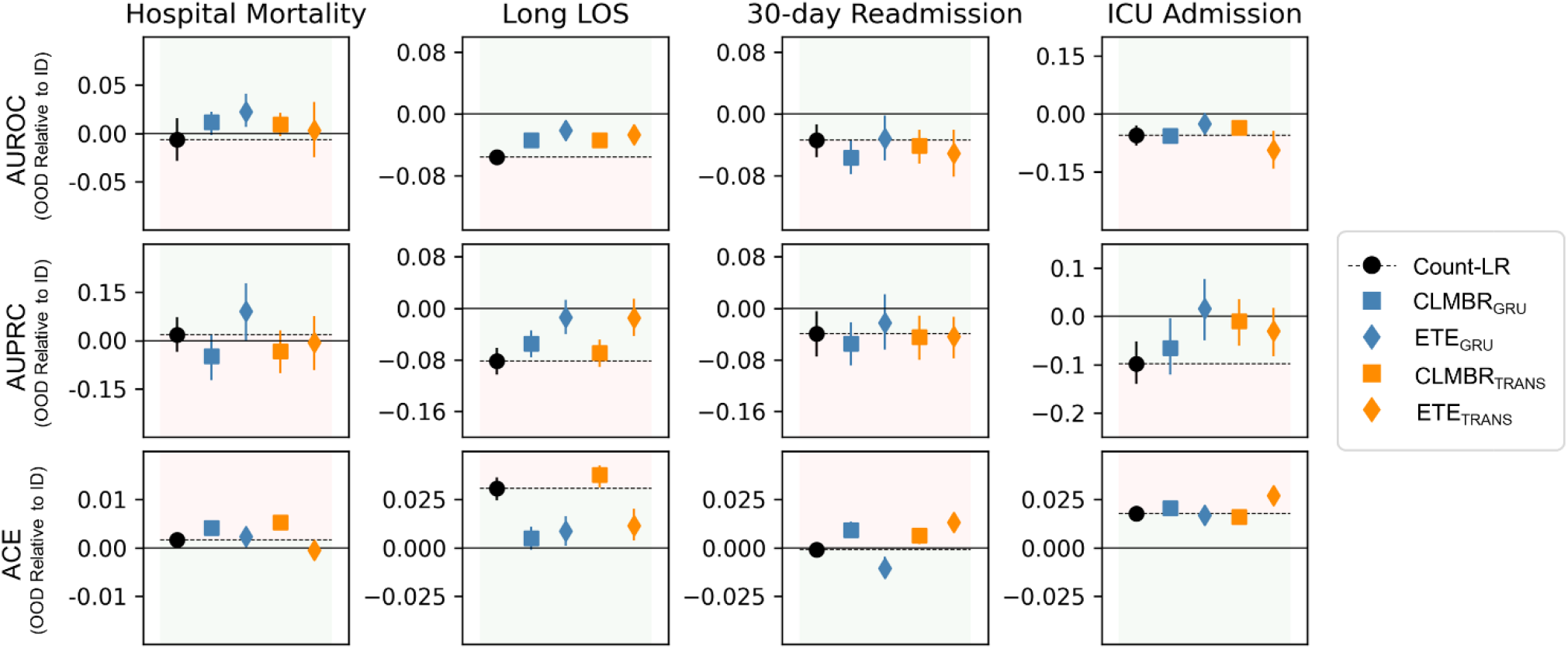
Relative out-of-distribution (OOD) performance (AUROC, AUPRC, and ACE) across representation construction and modeling approaches. Each relative performance was obtained by subtracting the model’s OOD performance in 2017-2021 (17-21) by its in-distribution (ID) performance in 2009-2012 (09-12) represented by the solid line. Colored regions indicate the range of values that is worse (red) or better (green) with respect to the relative OOD performance of logistic regression models trained on count-based representations. Error bars indicate 95% confidence interval obtained from 1000 bootstrap iterations. Supplementary Table 5 details the statistical results. Abbreviations: AUROC: area under the receiver operating characteristics curve; AUPRC: area under the precision recall curve; ACE: absolute calibration error; LOS: length of stay; ICU: intensive care unit; CLMBR: clinical language model-based representation; GRU: gated recurrent unit; Trans: transformer; LR: logistic regression; ETE: end-to-end.

Figure 5 plots the average binary cross entropy loss of GRU sequence models (trained using various hyperparameter settings) in the pretraining validation set against the average binary cross entropy loss of their downstream logistic regression models in each task and year group (see Supplementary Figure 1 for the same analysis conducted on CLMBR_TRANS_). The performance of GRU sequence models had high correlations (Pearson correlation coefficient) with the ID and OOD performance of their downstream logistic regression models in all tasks except for 30-day readmission.

**Figure 5.**
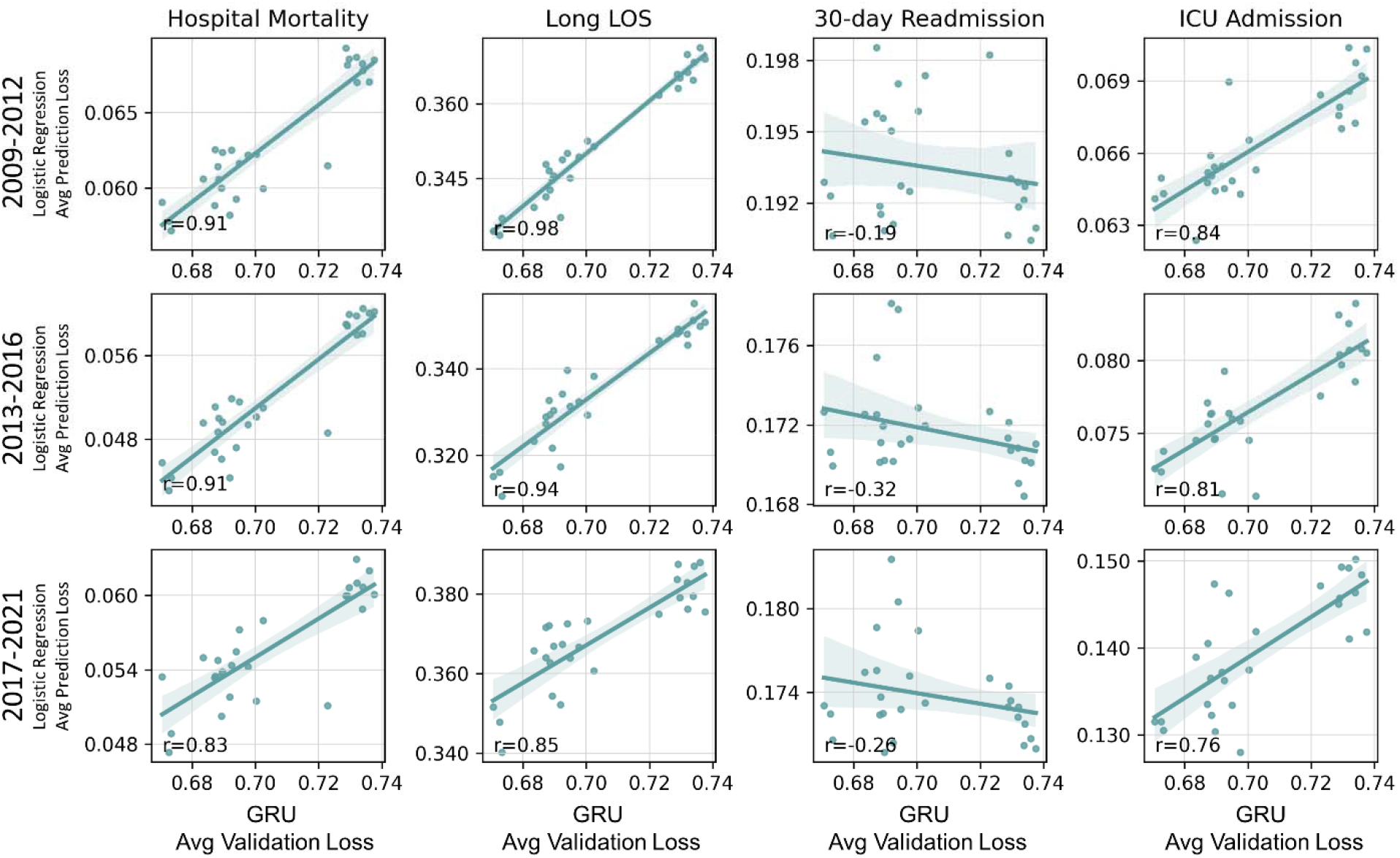
Correlation between the GRU sequence model’s validation performance and the performance of the downstream logistic regression models in each clinical prediction tasks. Performance for both the sequence model and the logistic regression model were measured using binary cross entropy loss. Each point in the scatter plot represents the GRU’s performance in the validation set and its downstream logistic regression model’s performance in the test set. Each GRU sequence model was pretrained using a different hyperparameter setting from the hyperparameter grid. Shaded error envelope represents the 95% confidence interval around the regression line. Abbreviations: GRU: gated recurrent unit; LOS: length of stay; ICU: intensive care unit.

Figure 6 plots the proportion of patients re-classified differently using CLMBR_GRU_ instead of count-LR in long LOS and ICU admission. There were generally more correct re-classifications than incorrect re-classifications across risk thresholds and year groups for long LOS. For ICU admission, there were more correct re-classifications in lower thresholds.

**Figure 6.**
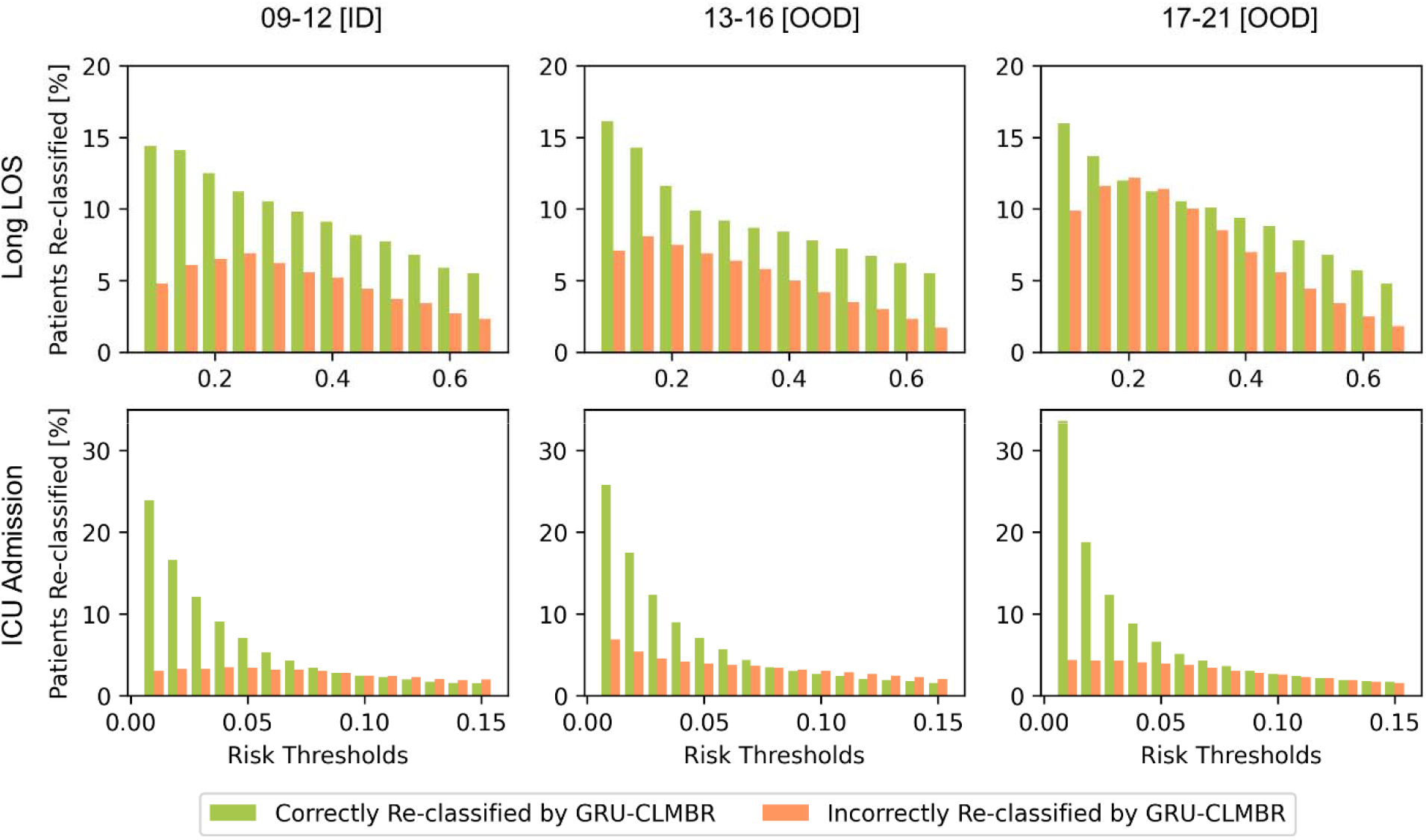
The proportion of patients correctly and incorrectly re-classified by CLMBR_GRU_. Green bars indicate the proportion of patients that were incorrectly classified by count-LR but were correctly re-classified by CLMBR_GRU_. Orange bars indicate the proportion of patients that were correctly classified by count-LR but were incorrectly re-classified by CLMBR_GRU_. Together, these represent the percentage of decisions that would have been affected if CLMBR_GRU_ models were used instead of count-LR. Abbreviations: count-LR: logistic regression models trained on count-based representations; CLMBR_GRU_: logistic regression models trained on gated recurrent unit-based CLMBR. CLMBR: clinical language model-based representations; LOS: length of stay; ICU: intensive care unit.

The sensitivity analysis where we trained and compared task-specific models on count-based representations and CLMBR using LightGBM instead of logistic regression showed qualitatively similar findings (Supplementary Experiment).

## DISCUSSION

We observed count-LR models resulted in large performance degradation over time for some tasks, namely long LOS and ICU admission. Models trained on CLMBR generally displayed better discrimination relative to count-LR in ID and OOD year groups but could result in worse OOD calibration. In addition, models trained on CLMBR often matched and were sometimes even better than their ETE counterparts. Finally, in general, CLMBR_GRU_ performed better than CLMBR_TRANS_, and its performance in the autoregressive sequence modeling task tracked closely with the ID and OOD performance of the downstream models for the majority of tasks considered.

Large-scale self-supervised pretraining takes place less frequently and enables machine learning practitioners to focus on rapid adaptation of these foundation models to downstream tasks. This research paper contributes more evidence that this approach brings not only performance benefits over traditional count-based models, but robustness benefits in the presence of temporal distribution shift. These benefits decrease the need for model retraining and preserves the clinical utility of models deployed into practice. The strong relationship between sequence modeling performance and the performance of downstream clinical prediction tasks suggests that the observed favorable ID and OOD performance can be directly attributed to the self-supervised pretraining. Whether the potential for worse calibration is clinically meaningful will depend on the specific use case.

The transformer architecture was more difficult to tune and performed worse than the GRU in both CLMBR-based models and ETE models. This may be explained by the smaller training set size used in this study, which totaled to ∼1.1 million patient days and ∼41.5 million medical codes across ∼29 thousand patients. In comparison, state of the art transformer-based natural language processing models leveraged much larger volumes of data, for example, ∼3,300 times larger (137 billion tokens) for BERT[11], and ∼53,000 times larger (2.2 trillion tokens) for RoBERTa[30].

We examined two attributes of OOD performance, namely absolute performance and relative (to ID) performance. It is notable that while CLMBR resulted in generally better absolute discrimination than count-LR, improvement in relative discrimination was more modest, with improvement only being observed for long LOS. It is likely that absolute performance is more meaningful to clinicians since better relative performance does not necessarily indicate better absolute performance[31]. However, decision makers may be more concerned about using a model that does not perform as well as that originally promised (relative performance). It is for that reason that we choose to report both aspects.

Despite the reasonable performance of CLMBR-based models in OOD year groups, they are not immune to the impact of distribution shift as seen in predicting ICU admissions and long LOS. Increasing parameter size and training set size, which have demonstrated benefits in other modalities[30, 32], could provide additional improvements to robustness. In addition, structure and certain types of invariances could be incorporated during pretraining or at the adaptation stage using metadata[33], regularization[34], or contrastive learning[35].

The strengths of this study include the evaluation of a novel approach to self-supervised representation learning on electronic health records, namely CLMBR, in both ID and OOD settings. Another strength is the adoption of interpretable metrics to evaluate the clinical impact of using CLMBR-based models instead of models trained on count-based representations. However, this study is limited as we only used a single dataset with a limited number of tasks. Performance of CLMBR may differ in other settings and with other asks. Another limitation is that we lack insight into scenarios in which CLMBR may be more or less helpful.

In conclusion, models trained on CLMBR generally displayed better discrimination relative to count-LR in both ID and OOD year groups. Models trained on CLMBR often matched or were better than their ETE counterparts. Finally, autoregressive sequence modeling performance tracked closely with the ID and OOD performance of the downstream models. These results suggest that pretraining EHR foundation models is a useful approach for developing clinical prediction models that perform well ID as well as OOD.

## Data Availability

The Stanford Medicine Research Data Repository is not made publicly available. The code for all analyses is open-source and available at https://github.com/som-shahlab/temp_ds_shift_robustness

## Ethics approval and consent to participate

This study used de-identified data and so the requirement for Institutional Review Board approval and participant informed consent were waived by Stanford Medical Center.

## Acknowledgments

Not applicable.

## Authors’ contributions

LLG, JF, and LS conceptualized and designed the study with input from all authors. L.L.G. performed all experiments. ES and SRP contributed to the codebase. LLG, JF, and LS analyzed and interpreted results with input from all authors. LLG wrote the manuscript. All authors revised and commented on the manuscript. All authors read and approved the final manuscript.

