## Supplementary for "EHR Foundation Models Improve Robustness in the Presence of Temporal Distribution Shift"

**Supplementary Content**

| Pages |  |  |
| --- | --- | --- |
| 2-6 | **Supplementary Methods** | Additional details on model hyperparameters |
| 7 | **Supplementary Table 1** | Cohort characteristics by year and outcome prevalence |
| 8 | **Supplementary Table 2** | Differences in in-distribution performance in 2009-2012 across representation construction and modeling approaches |
| 9 | **Supplementary Table 3** | Differences in out-of-distribution performance in 2017-2021 across representation construction and modeling approaches |
| 10 | **Supplementary Table 4** | Difference in relative performance in 2017-2021 across representation construction and modeling approaches |
| 11 | **Supplementary**  **Figure 1** | Correlation between the transformer model’s validation performance and the performance of the downstream logistic regression models in each clinical prediction tasks. |
| 12-16 | **Supplementary Experiment** | Additional experiment in which LightGBM models were used as adapter models instead of Logistic Regression |

**Supplementary Methods.** Additional details on clinical outcomes, feature extraction, models and learning algorithms.

I. GRU and Transformer hyperparameters
II. Selected hyperparameter settings for CLMBR and end-to-end models
III. Selected hyperparameter setting for logistic regression models

**Supplementary Methods I. GRU and transformer hyperparameters**

| **Architecture** | **Hyperparameter** | **Values** |
| --- | --- | --- |
| GRU ^a^ | Learning rate | 0.01, 0.001, 0.0001 |
| GRU | Dropout | 0, 0.1, 0.2 |
| GRU | L2 | 0.1, 0.01, 0.001 |
| Transformer | Learning rate | 0.0001, 0.00001 |
| Transformer | Dropout | 0, 0.2, 0.4 |
| Transformer | L2 | 0.1, 0.01 |
| Transformer | Number of layers ^b^ | 6, 12 |
| Transformer | Code dropout | 0.2, 0.4 |

GRU and transformer models were trained using a batch size of 2000 for 200 epochs. End-to-end models were trained for 50 epochs.

^a^ All GRU architectures consisted of one layer. Code dropout was set at 0.2.

^b^ Each layer in the transformer architecture consisted of a multi-head attention (number of heads = 8) layer and two feed-forward layers.

Abbreviations: CLMBR: clinical language model-based representations; GRU: gated recurrent unit.

**Supplementary Methods II**. **Selected model hyperparameters for CLMBR and end-to-end models**

| **Architecture** | **Task** | **Hyperparameter** | **Value** |
| --- | --- | --- | --- |
| GRU | Sequence modeling | Learning rate | 0.01 |
| GRU | Sequence modeling | Dropout | 0.1 |
| GRU | Sequence modeling | L2 | 0.1 |
| Transformer | Sequence modeling | Learning rate | 0.0001 |
| Transformer | Sequence modeling | Dropout | 0.4 |
| Transformer | Sequence modeling | L2 | 0.01 |
| Transformer | Sequence modeling | Number of layers | 6 |
| Transformer | Sequence modeling | Code dropout | 0.2 |
| GRU | Hospital Mortality | Learning rate | 0.01 |
| GRU | Hospital Mortality | Dropout | 0.2 |
| GRU | Hospital Mortality | L2 | 0.1 |
| Transformer | Hospital Mortality | Learning rate | 0.0001 |
| Transformer | Hospital Mortality | Dropout | 0 |
| Transformer | Hospital Mortality | L2 | 0.01 |
| Transformer | Hospital Mortality | Number of layers | 6 |
| Transformer | Hospital Mortality | Code dropout | 0.4 |
| GRU | Long LOS | Learning rate | 0.001 |
| GRU | Long LOS | Dropout | 0 |
| GRU | Long LOS | L2 | 0.01 |
| Transformer | Long LOS | Learning rate | 0.0001 |
| Transformer | Long LOS | Dropout | 0 |
| Transformer | Long LOS | L2 | 0.01 |
| Transformer | Long LOS | Number of layers | 6 |
| Transformer | Long LOS | Code dropout | 0.4 |
| GRU | ICU Admission | Learning rate | 0.001 |
| GRU | ICU Admission | Dropout | 0 |
| GRU | ICU Admission | L2 | 0.01 |
| Transformer | ICU Admission | Learning rate | 0.0001 |
| Transformer | ICU Admission | Dropout | 0.2 |
| Transformer | ICU Admission | L2 | 0.1 |
| Transformer | ICU Admission | Number of layers | 6 |
| Transformer | ICU Admission | Code dropout | 0.2 |
| GRU | 30d readmission | Learning rate | 0.0001 |
| GRU | 30d readmission | Dropout | 0 |
| GRU | 30d readmission | L2 | 0.1 |
| Transformer | 30d readmission | Learning rate | 0.0001 |
| Transformer | 30d readmission | Dropout | 0 |
| Transformer | 30d readmission | L2 | 0.01 |
| Transformer | 30d readmission | Number of layers | 6 |
| Transformer | 30d readmission | Code dropout | 0.4 |

Abbreviations: CLMBR: clinical language model-based representations; GRU: gated recurrent unit; ICU: intensive care unit; LOS: length of stay

**Supplementary Methods III**. **Selected model hyperparameters for logistic regression**

| **Featurization** | **Task** | **Hyperparameter^a^ Value** |
| --- | --- | --- |
| Count-based | Hospital Mortality | 0.01 |
| Count-based | Long LOS | 0.01 |
| Count-based | ICU Admission | 0.01 |
| Count-based | 30d readmission | 0.001 |
| GRU-CLMBR | Hospital Mortality | 0.1 |
| GRU-CLMBR | Long LOS | 0.01 |
| GRU-CLMBR | ICU Admission | 0.1 |
| GRU-CLMBR | 30d readmission | 0.01 |
| Transformer-CLMBR | Hospital Mortality | 0.001 |
| Transformer-CLMBR | Long LOS | 0.001 |
| Transformer-CLMBR | ICU Admission | 0.01 |
| Transformer-CLMBR | 30d readmission | 0.001 |

^a^Hyperparameter for logistic regression is the inverse of L2 regularization strength, and thus smaller values indicate stronger regularization. Search was conducted over values ranging from 10^-6^ to 10^2^ in powers of 10

Abbreviations: CLMBR: clinical language model-based representations; GRU: gated recurrent unit; ICU: intensive care unit; LOS: length of stay

**Supplementary Table 1. Cohort characteristics by year**

|  | **Year** | | | | | | | | | | | | |
| --- | --- | --- | --- | --- | --- | --- | --- | --- | --- | --- | --- | --- | --- |
|  | **2009** | **2010** | **2011** | **2012** | **2013** | **2014** | **2015** | **2016** | **2017** | **2018** | **2019** | **2020** | **2021** |
| **No.** | 12727 | 12930 | 13104 | 13136 | 13117 | 15235 | 17241 | 16980 | 17198 | 21187 | 22514 | 22352 | 13509 |
| **Mean age in years ± SD** | 57±18 | 57±18 | 57±18 | 57±18 | 57±18 | 53±19 | 52±19 | 52±19 | 53±19 | 53±20 | 53±20 | 53±20 | 54±20 |
| **Sex, No. (%)** | | | | | | | | | | | | | |
| **Female** | 6572 (52%) | 6579 (51%) | 6767 (52%) | 6576 (50%) | 6668 (51%) | 8820 (58%) | 10483 (61%) | 10310 (61%) | 10269 (60%) | 12874 (61%) | 13556 (60%) | 13234 (59%) | 8010 (59%) |
| **Male** | 6154 (48%) | 6351 (49%) | 6337 (48%) | 6559 (50%) | 6448 (49%) | 6414 (42%) | 6757 (39%) | 6670 (39%) | 6929 (40%) | 8313 (39%) | 8956 (40%) | 9116 (41%) | 5496 (41%) |
| **Race, No. (%)** | | | | | | | | | | | | | |
| **White** | 7054 (55%) | 8106 (63%) | 8091 (62%) | 7848 (60%) | 7585 (58%) | 8092 (53%) | 8620 (50%) | 8174 (48%) | 8328 (48%) | 10164 (48%) | 10866 (48%) | 10101 (45%) | 5908 (44%) |
| **Other** | 5673 (45%) | 4824 (37%) | 5013 (38%) | 5288 (40%) | 5532 (42%) | 7143 (47%) | 8621 (50%) | 8806 (52%) | 8870 (52%) | 11023 (52%) | 11648 (52%) | 12251 (55%) | 7601 (56%) |
| **Clinical Outcome, No. (%)** | | | | | | | | | | | | | |
| **In-Hospital Mortality** | 267 (2%) | 317 (2%) | 337 (3%) | 337 (3%) | 340 (3%) | 321 (2%) | 345 (2%) | 345 (2%) | 335 (2%) | 402 (2%) | 379 (2%) | 459 (2%) | 249 (2%) |
| **LOS >7 Days** | 2680 (21%) | 2718 (21%) | 2719 (21%) | 2739 (21%) | 2703 (21%) | 2939 (19%) | 3065 (18%) | 3364 (20%) | 3263 (19%) | 3767 (18%) | 4078 (18%) | 4300 (19%) | 2709 (20%) |
| **30-Day Readmission** | 658 (5%) | 760 (6%) | 706 (6%) | 681 (5%) | 687 (5%) | 780 (5%) | 813 (5%) | 767 (5%) | 762 (5%) | 1012 (5%) | 1079 (5%) | 1080 (5%) | 619 (5%) |
| **ICU Admission** | 116 (1%) | 315 (2%) | 327 (2%) | 325 (2%) | 356 (3%) | 390 (3%) | 393 (2%) | 481 (3%) | 428 (2%) | 613 (3%) | 704 (3%) | 1116 (5%) | 591 (4%) |

Abbreviations. SD: standard deviation; LOS: long length of stay; ICU: intensive care unit.

**Supplementary Table 2. Differences in in-distribution performance in 2009-2012 across representation construction and modeling approaches.**

|  |  | Median Difference (Lower 95% CI, Upper 95% CI)^a^ | | | | |
| --- | --- | --- | --- | --- | --- | --- |
|  |  | Comparison of models trained on CLMBR with models trained on count-based representations | | Comparison of models trained on CLMBR with end-to-end models using the same architecture | | Comparison of GRU and trans architecture for CLMBR |
| Task | Metric | GRU-CLMBR  vs.  Counts | Trans-CLMBR  vs.  Counts | GRU-CLMBR  vs.  GRU-ETE | Trans-CLMBR  vs. Trans-ETE | GRU-CLMBR  vs.  Trans-CLMBR |
| Hospital  Mortality | AUC | **0.058 (0.041,0.077)** | **0.049 (0.034,0.064)** | 0.006  (-0.005,0.016) | **0.031 (0.015,0.051)** | **0.009 (0.001,0.018)** |
|  | AUPRC | **0.245 (0.174,0.314)** | **0.157 (0.105,0.21)** | 0.017  (-0.046,0.08) | 0.053  (-0.01,0.116) | **0.088 (0.024,0.148)** |
|  | ACE | 0.001  (-0.001,0.003) | 0.0  (-0.002,0.002) | **-0.004**  **(-0.006,-0.003)** | **-0.005**  **(-0.008,-0.002)** | 0.001  (-0.002,0.003) |
| Long  LOS | AUC | **0.057 (0.049,0.066)** | **0.038 (0.03,0.047)** | -0.002  (-0.007,0.004) | -0.001  (-0.008,0.006) | **0.019 (0.013,0.025)** |
|  | AUPRC | **0.138 (0.118,0.16)** | **0.086 (0.066,0.106)** | 0.002  (-0.014,0.019) | **0.024 (0.004,0.043)** | **0.052 (0.035,0.068)** |
|  | ACE | 0.005  (-0.001,0.01) | 0.001  (-0.004,0.005) | **-0.015**  **(-0.023,-0.005)** | **-0.029**  **(-0.035,-0.02)** | 0.005  (-0.001,0.009) |
| 30 Day  Readmission | AUC | 0.004  (-0.011,0.02) | 0.004  (-0.012,0.02) | **0.017 (0.003,0.032)** | **0.024 (0.006,0.04)** | 0.0  (-0.014,0.013) |
|  | AUPRC | -0.022  (-0.052,0.006) | -0.014  (-0.042,0.016) | **0.033 (0.009,0.059)** | **0.059 (0.032,0.087)** | -0.008  (-0.035,0.017) |
|  | ACE | -0.004  (-0.008,0.001) | **-0.004**  **(-0.007,-0.0)** | **-0.009**  **(-0.01,-0.005)** | **-0.002**  **(-0.003,-0.0)** | 0.0  (-0.002,0.002) |
| ICU  Admission | AUC | **0.074 (0.05,0.096)** | **0.061 (0.038,0.082)** | **0.041 (0.025,0.059)** | **0.155 (0.115,0.196)** | **0.013 (0.001,0.025)** |
|  | AUPRC | **0.149 (0.077,0.217)** | 0.042  (-0.018,0.103) | **0.138 (0.083,0.199)** | **0.106 (0.053,0.161)** | **0.106 (0.048,0.161)** |
|  | ACE | -0.002  (-0.005,0.001) | -0.001  (-0.005,0.002) | **-0.006**  **(-0.008,-0.003)** | **-0.004**  **(-0.006,-0.002)** | -0.001  (-0.002,0.001) |

^a^ Confidence intervals were calculated from the distribution of differences obtained over 10,000 bootstrap iterations. Bolded values indicate statistically significant increase in performance. Italicized values indicate statistically significant decrease in performance.

Abbreviations: CI: confidence interval; ACE: absolute calibration error; GRU: gated recurrent unit; trans: transformer; CLMBR: clinical language model-based representation; ETE: end-to-end; LOS: length of stay; ICU: intensive care unit

**Supplementary Table 3. Differences in out-of-distribution performance in 2017-2021 across representation construction and modeling approaches.**

|  |  | Median Difference (Lower 95% CI, Upper 95% CI) ^a^ | | | | |
| --- | --- | --- | --- | --- | --- | --- |
|  |  | Comparison of models trained on CLMBR with models trained on count-based representations | | Comparison of models trained on CLMBR with end-to-end models using the same architecture | | Comparison of GRU and trans architecture for CLMBR |
| Task | Metric | GRU-CLMBR  vs  Counts | Trans-CLMBR  vs  Counts | GRU-CLMBR  vs  GRU-ETE | Trans-CLMBR  vs Trans-ETE | GRU-CLMBR  vs  Trans-CLMBR |
| Hospital  Mortality | AUC | **0.075 (0.059,0.092)** | **0.065 (0.048,0.083)** | -0.007  (-0.014,0.002) | **0.036 (0.022,0.051)** | **0.01 (0.004,0.018)** |
|  | AUPRC | **0.179 (0.122,0.234)** | **0.104 (0.06,0.154)** | *-0.122*  *(-0.176,-0.064)* | 0.023  (-0.024,0.072) | **0.074 (0.023,0.122)** |
|  | ACE | *0.005 (0.002,0.007)* | *0.006 (0.003,0.008)* | **-0.002**  **(-0.003,-0.001)** | *0.004 (0.002,0.006)* | -0.001  (-0.002,0.0) |
| Long  LOS | AUC | **0.08 (0.07,0.089)** | **0.06 (0.049,0.07)** | *-0.015*  *(-0.02,-0.01)* | *-0.009*  *(-0.015,-0.003)* | **0.02 (0.014,0.025)** |
|  | AUPRC | **0.166 (0.147,0.184)** | **0.099 (0.08,0.118)** | *-0.04*  *(-0.052,-0.028)* | *-0.032*  *(-0.047,-0.014)* | **0.067 (0.053,0.08)** |
|  | ACE | **-0.021**  **(-0.025,-0.017)** | *0.007 (0.003,0.012)* | **-0.018**  **(-0.023,-0.013)** | **-0.003**  **(-0.007,0.001)** | **-0.028**  **(-0.031,-0.025)** |
| 30 Day  Readmission | AUC | -0.018  (-0.037,0.001) | -0.003  (-0.017,0.01) | -0.007  (-0.026,0.01) | **0.033 (0.017,0.048)** | -0.015  (-0.033,0.003) |
|  | AUPRC | -0.036  (-0.06,-0.013) | -0.019  (-0.037,0.002) | 0.001  (-0.02,0.025) | **0.057 (0.037,0.078)** | -0.018  (-0.039,0.003) |
|  | ACE | *0.006 (0.004,0.009)* | *0.003 (0.0,0.007)* | *0.012 (0.01,0.014)* | **-0.009**  **(-0.01,-0.008)** | *0.003 (0.002,0.004)* |
| ICU  Admission | AUC | **0.073 (0.054,0.091)** | **0.082 (0.063,0.101)** | 0.009  (-0.004,0.021) | **0.215 (0.191,0.239)** | -0.009  (-0.02,0.003) |
|  | AUPRC | **0.18 (0.143,0.216)** | **0.128 (0.095,0.164)** | **0.058 (0.017,0.098)** | **0.126 (0.086,0.164)** | **0.052 (0.01,0.091)** |
|  | ACE | 0.001  (-0.0,0.001) | **-0.003**  **(-0.004,-0.002)** | **-0.002**  **(-0.003,-0.001)** | **-0.015 (**  **-0.016,-0.014)** | *0.004 (0.003,0.005)* |

^a^ Confidence intervals were calculated from the distribution of differences obtained over 10,000 bootstrap iterations. Bolded values indicate statistically significant increase in performance. Italicized values indicate statistically significant decrease in performance.

Abbreviations: CI: confidence interval; ACE: absolute calibration error; GRU: gated recurrent unit; trans: transformer; CLMBR: clinical language model-based representation; ETE: end-to-end; LOS: length of stay; ICU: intensive care unit

**Supplementary Table 4**. **Differences in relative performance of 2017-2021 across representation construction and modeling approaches.**

|  |  | Median Difference (Lower 95% CI, Upper 95% CI) | | | | |
| --- | --- | --- | --- | --- | --- | --- |
|  |  | Comparison of models trained on CLMBR with models trained on count-based representations | | Comparison of models trained on CLMBR with end-to-end models using the same architecture | | Comparison of GRU and Trans architecture for CLMBR |
| Task | Metric | GRU-CLMBR  Minus  Counts | Trans-CLMBR  Minus  Counts | GRU-CLMBR  Minus  GRU-ETE | Trans-CLMBR  Minus Trans-ETE | GRU-CLMBR  Minus  Trans-CLMBR |
| Hospital  Mortality | AUC | 0.017  (-0.008,0.044) | 0.015  (-0.007,0.039) | -0.012  (-0.025,0.001) | 0.005  (-0.018,0.028) | 0.002  (-0.01,0.013) |
|  | AUPRC | -0.066  (-0.16,0.035) | -0.052  (-0.126,0.02) | *-0.142*  *(-0.225,-0.063)* | -0.027  (-0.104,0.051) | -0.017  (-0.093,0.063) |
|  | ACE | *0.004*  *(0.001,0.007)* | *0.005*  *(0.003,0.008)* | *0.002*  *(0.0,0.005)* | *0.009*  *(0.005,0.012)* | -0.002  (-0.004,0.001) |
| Long  LOS | AUC | **0.022 (0.01,0.035)** | **0.022 (0.009,0.034)** | *-0.013*  *(-0.021,-0.005)* | -0.008  (-0.018,0.002) | 0.001  (-0.007,0.01) |
|  | AUPRC | 0.028  (-0.001,0.057) | 0.012  (-0.016,0.042) | -0.041  (-0.062,-0.021) | -0.055  (-0.081,-0.028) | 0.016  (-0.007,0.039) |
|  | ACE | **-0.026**  **(-0.033,-0.019)** | *0.006 (0.0,0.013)* | -0.004  (-0.016,0.007) | *0.025 (0.017,0.034)* | **-0.033**  **(-0.038,-0.027)** |
| 30 Day  Readmission | AUC | -0.022  (-0.045,0.002) | -0.008  (-0.028,0.013) | *-0.024*  *(-0.045,-0.001)* | 0.008  (-0.014,0.032) | -0.014  (-0.036,0.009) |
|  | AUPRC | -0.014  (-0.053,0.024) | -0.004  (-0.041,0.028) | -0.032  (-0.063,0.002) | -0.001  (-0.036,0.034) | -0.008  (-0.044,0.025) |
|  | ACE | *0.01 (0.004,0.016)* | *0.007 (0.003,0.012)* | *0.02 (0.017,0.023)* | **-0.007**  **(-0.009,-0.005)** | *0.003 (0.0,0.006)* |
| ICU  Admission | AUC | -0.001  (-0.033,0.028) | 0.021  (-0.008,0.048) | *-0.034*  *(-0.055,-0.013)* | **0.06 (0.012,0.104)** | *-0.022*  *(-0.039,-0.006)* |
|  | AUPRC | 0.036  (-0.042,0.107) | **0.087 (0.022,0.155)** | *-0.081*  *(-0.154,-0.006)* | 0.02  (-0.052,0.084) | -0.055  (-0.125,0.013) |
|  | ACE | 0.003  (-0.001,0.006) | -0.002  (-0.006,0.002) | *0.004 (0.0,0.007)* | **-0.011**  **(-0.013,-0.009)** | *0.004 (0.002,0.006)* |

Each relative performance was obtained by subtracting the model’s OOD performance in 2017-2021 by its in-distribution performance in 2009-2012. Confidence intervals were calculated from the distribution of differences obtained over 1000 bootstrap iterations. Bolded values indicate significant increase in performance. Italicized values indicate significant decrease in performance.

CI: confidence interval; ACE: absolute calibration error; GRU: gated recurrent unit; Trans: transformer; CLMBR: clinical language model-based representation; ETE: end-to-end; LOS: length of stay; OOD: out-of-distribution.

**Supplementary Figure 1**. Correlation between the transformer model’s validation performance and the performance of the downstream logistic regression models in each clinical prediction tasks. Performance for both the transformer and the logistic regression models were measured using binary cross entropy loss. Shaded error envelope represents the 95% confidence interval around the regression line. Note that the correlations are inflated (especially in 30-day readmission) by a cluster of transformer models that performed poorly in the sequence modeling task due to suboptimal hyperparameter settings. These transformer models also produced poorly performing downstream task-specific models (example highlighted by the blue rectangle in the top left panel). We observed that the performance of transformer models tend to be more sensitive to changes in hyperparameter settings, for example in the learning rate.

Abbreviations: LOS: length of stay; ICU: intensive care unit.

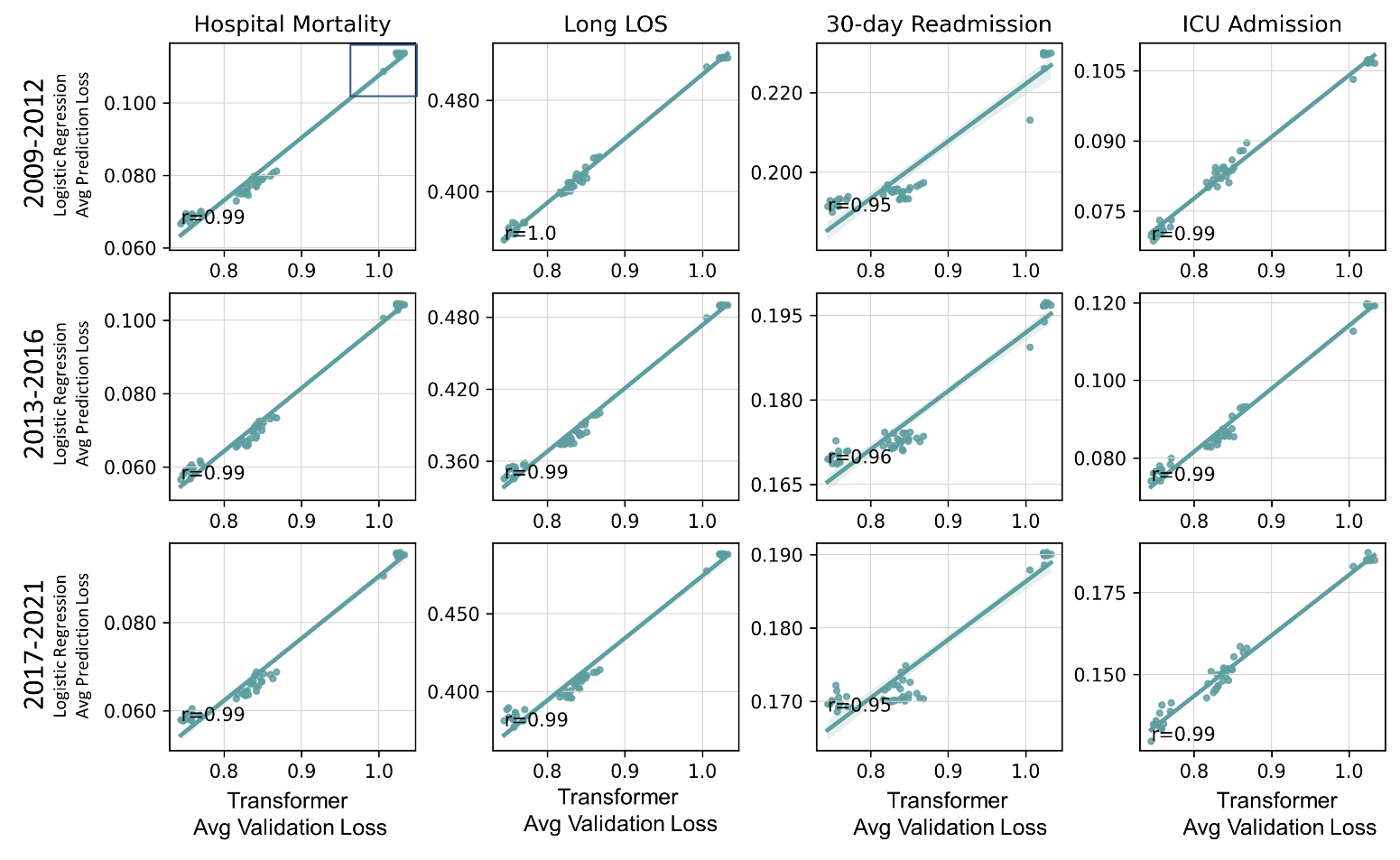

**Supplementary Experiment.** Additional experiment in which LightGBM models were used as clinical prediction models instead of Logistic Regression.

I. Selected LightGBM hyperparameter values
II. The impact of temporal dataset shift on the performance of LightGBM models trained on count-based representations
III. Performance across representation construction and modeling approaches in each year group
IV. Relative out-of-distribution performance across representation construction and modeling approaches

**Supplementary Experiment I**. Selected light gradient boosting machine (LightGBM) hyperparameter values

| **Featurization** | **Task** | **Hyperparameter^a^ Values** |
| --- | --- | --- |
| Count-based | Hospital Mortality | **lr**: 0.01; **num_leaves**: 100; **boosting_type**: goss; **n_estimators**: 312 |
| Count-based | Long LOS | **lr**: 0.01; **num_leaves**: 100; **boosting_type**: goss; **n_estimators**: 312 |
| Count-based | ICU Admission | **lr**: 0.01; **num_leaves**: 100; **boosting_type**: gbdt; **n_estimators**: 650 |
| Count-based | 30d readmission | **lr**: 0.01; **num_leaves**: 100; **boosting_type**: goss; **n_estimators**: 371 |
| GRU-CLMBR | Hospital Mortality | **lr**: 0.01; **num_leaves**: 100; **boosting_type**: goss; **n_estimators**: 285 |
| GRU-CLMBR | Long LOS | **lr**: 0.01; **num_leaves**: 100; **boosting_type**: gbdt; **n_estimators**: 769 |
| GRU-CLMBR | ICU Admission | **lr**: 0.01; **num_leaves**: 100; **boosting_type**: goss; **n_estimators**: 249 |
| GRU-CLMBR | 30d readmission | **lr**: 0.01; **num_leaves**: 100; **boosting_type**: goss; **n_estimators**: 254 |
| Transformer-CLMBR | Hospital Mortality | **lr**: 0.01; **num_leaves**: 100; **boosting_type**: goss; **n_estimators**: 233 |
| Transformer-CLMBR | Long LOS | **lr**: 0.01; **num_leaves**: 100; **boosting_type**: gbdt; **n_estimators**: 544 |
| Transformer-CLMBR | ICU Admission | **lr**: 0.01; **num_leaves**: 100; **boosting_type**: goss; **n_estimators**: 254 |
| Transformer-CLMBR | 30d readmission | **lr**: 0.01; **num_leaves**: 100; **boosting_type**: goss; **n_estimators**: 320 |

^a^Grid search was conducted over learning rate (“lr”: 0.1, 0.2, 0.01), number of leaves (“num_leaves”: 100, 300), and boosting type (“boosting_type”: gbdt, dart, goss). The number of trees (“n_estimators”) for gbdt and goss was determined based on performance in the validation set via early stopping. The number of trees for dart was set at 1000.

**Supplementary Experiment II**. The impact of temporal dataset shift on the performance of LightGBM models trained on count-based representations. Shaded regions indicate time windows in which performance in out-of-distribution years (2013 - 2021) is worse (red) or better (green) than performance in the in-distribution year group (2009-2012). Oracle models were trained and evaluated on each of the out-of-distribution years. Error bars indicate 95% confidence interval obtained from 1000 bootstrap iterations.

Abbreviations: AUROC: area under the receiver operating characteristics curve; AUPRC: area under the precision recall curve; ACE: absolute calibration error; LOS: length of stay; ICU: intensive care unit.

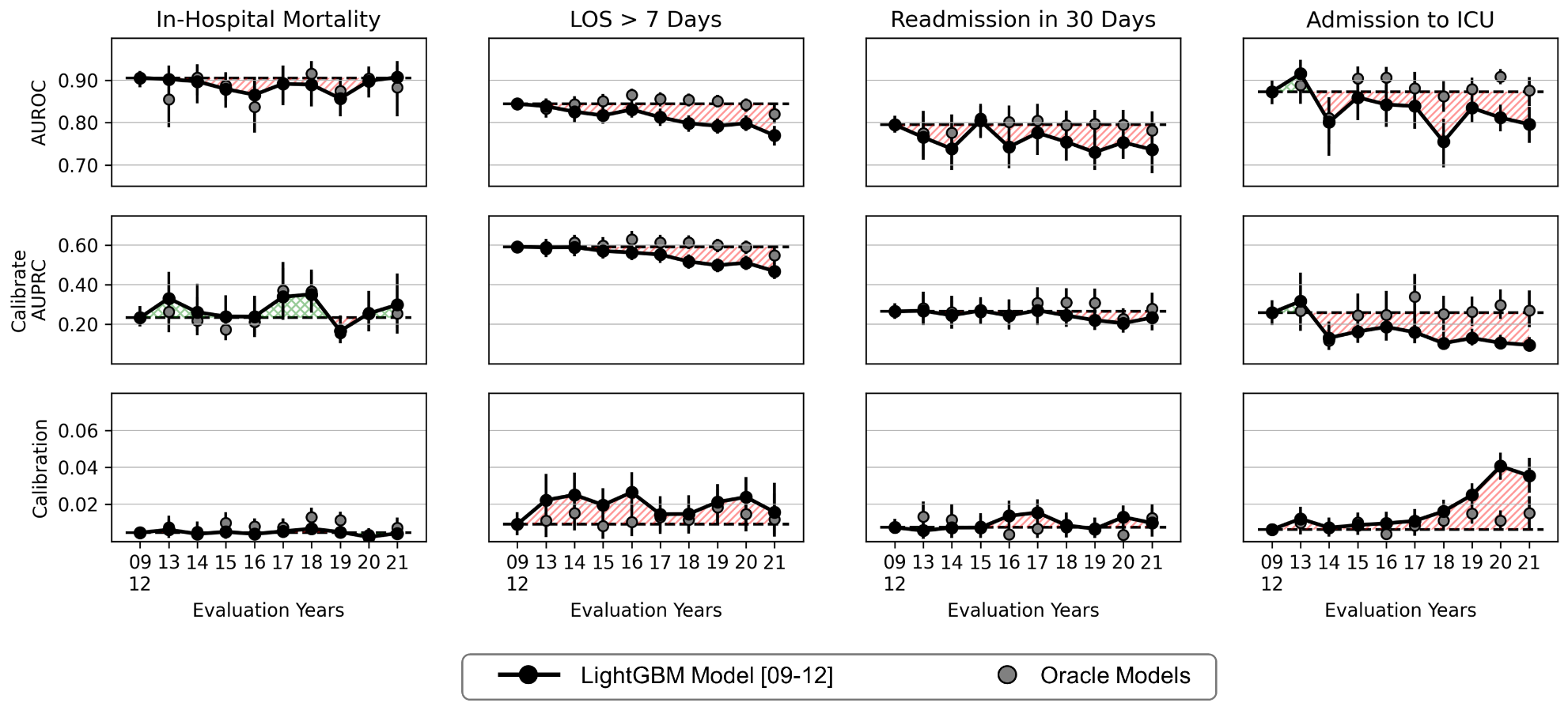

**Supplementary Experiment III**. Relative performance (AUROC, AUPRC and calibration measured using ACE) to count-GBM across featurization and modeling approaches in each year group, in which 2009-2012 (09-12) is the in-distribution year group and 2013-2016 (13-16) and 2017-2021(17-21) are the out-of-distribution year groups. Note that GRU-ETE and Trans-ETE results are the same as the main manuscript figure and are displayed here for comparison. Colored regions indicate the range of performance that is worse (red) or better (green) with respect to logistic regression models trained on count-based representations. Squares are CLMBR-based models and diamonds are ETE models. Error bars indicate 95% confidence interval obtained from 1000 bootstrap iterations.

Abbreviations: AUROC: area under the receiver operating characteristics curve; AUPRC: area under the precision recall curve; ACE: absolute calibration error; LOS: length of stay; ICU: intensive care unit; GBM: light gradient boosting machine; CLMBR: clinical language model-based representation; Trans: Transformer; ETE: end-to-end.

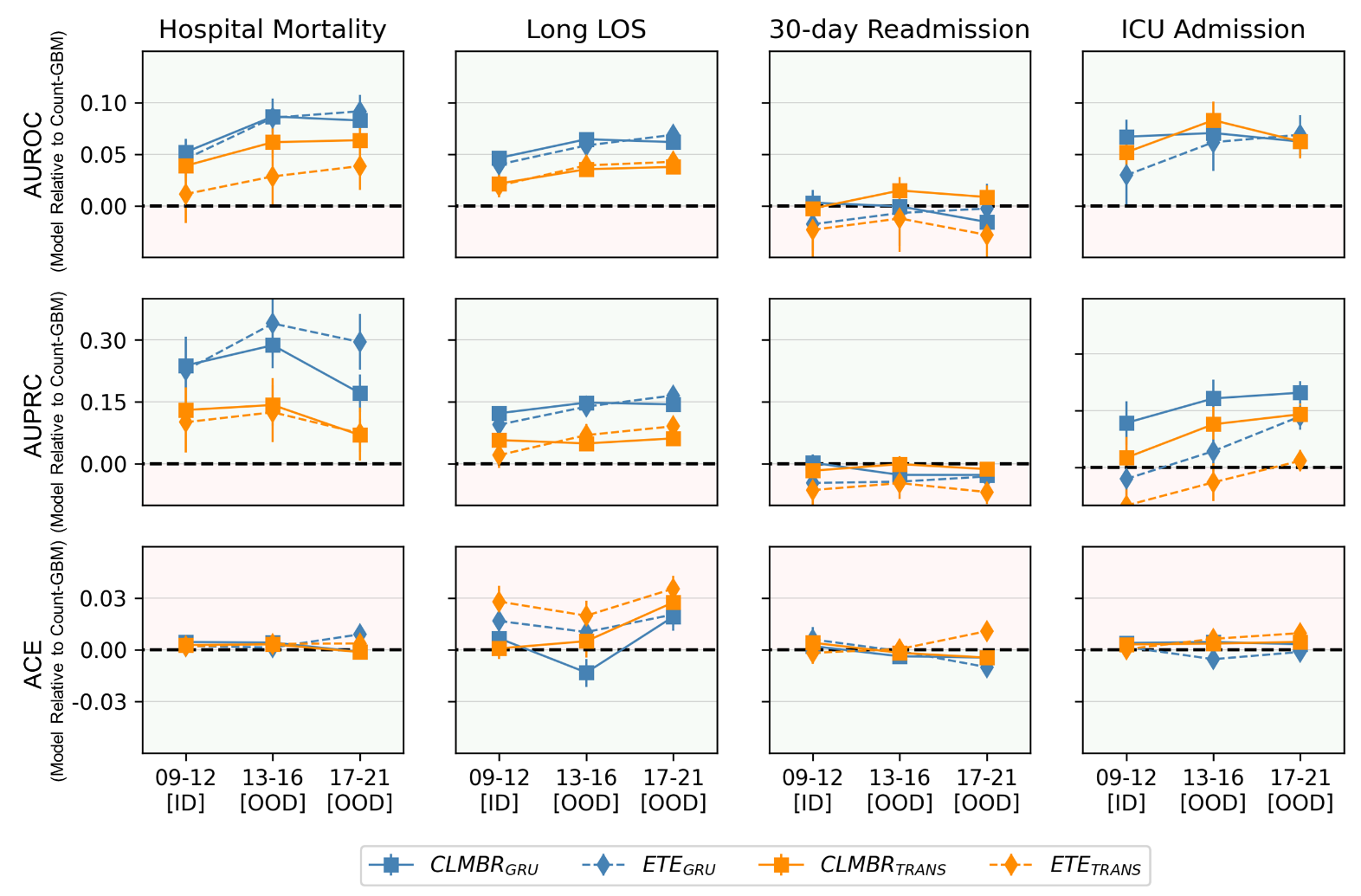

**Supplementary Experiment IV**. Relative out-of-distribution (OOD) performance (AUROC, AUPRC and calibration measured using ACE) across featurization and modeling approaches. Each relative performance was obtained by subtracting the model’s OOD performance in 2017-2021 by its in-distribution (ID) performance in 2009-2012 represented by the solid line. Note that GRU-ETE and Trans-ETE results are the same as the main manuscript figure and are displayed here for comparison. Colored regions indicate the range of performance that is worse (red) or better (green) with respect to the relative OOD performance of light GBM models trained on count-based representations. Error bars indicate 95% confidence interval obtained from 1000 bootstrap iterations.

Abbreviations: AUROC: area under the receiver operating characteristics curve; AUPRC: area under the precision recall curve; ACE: absolute calibration error; LOS: length of stay; ICU: intensive care unit; GBM: light gradient boosting machines; CLMBR: clinical language model-based representation; Trans: Transformer; ETE: end-to-end.

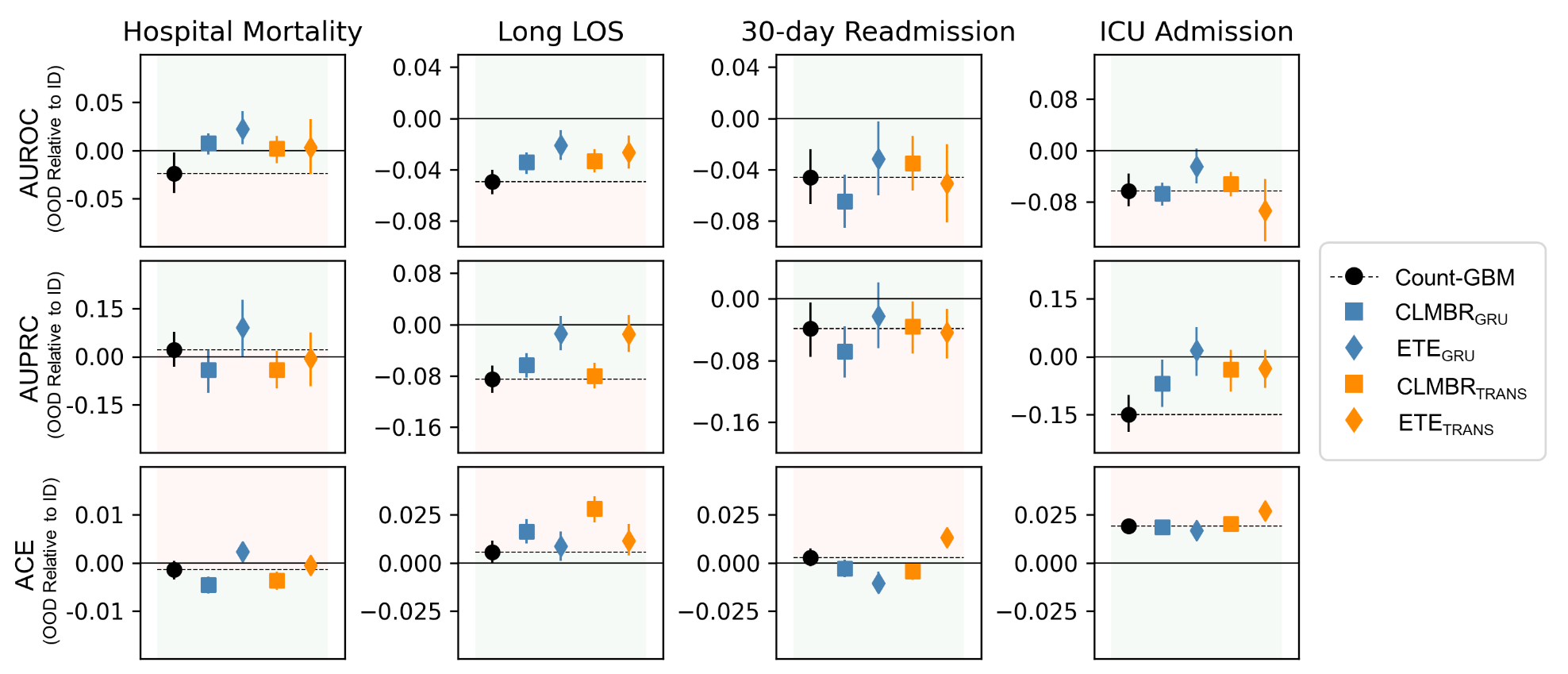
